# Wearable vibrotactile stimulation shirts and gloves for upper extremity stroke rehabilitation: A pilot randomized controlled trial

**DOI:** 10.64898/2026.07.26.26358522

**Authors:** Walaa Ayyad, Yeongdae Kim, Nathan Odom, Mazen Al Borno

## Abstract

**Objective:** Conduct a randomized controlled trial to investigate the safety, feasibility, and efficacy of wearable vibrotactile stimulation shirts and gloves for upper extremity stroke rehabilitation at the inpatient rehabilitation unit. The primary outcome measure was the Fugl-Meyer Assessment Upper Extremity (FMA-UE) motor score. The secondary outcome measures were the Modified Ashworth Scale (MAS) and statistics on device safety and feasibility. Exploratory outcome measures were the FMA-UE sensation, passive joint motion, and joint pain scores. Also, the MAS for the fingers, wrist, and elbow scores.

**Methods:** A total of 24 ischemic stroke patients were recruited for this study (45 to 85 years old) during their stay at the inpatient rehabilitation unit, which averaged 15.3 ± 4.5 days. Patients were randomly assigned to a treatment or control group, with 12 participants in each group. The control group received conventional therapy only, while the treatment group received both conventional therapy and vibrotactile stimulation. Participants in the treatment group wore the vibrotactile stimulation gloves and shirts for 1.5 hours per day, 5 days per week.

Upper extremity impairment and spasticity were assessed with the FMA-UE and MAS at both admission and discharge from the rehabilitation unit. In addition, feedback from patients in the treatment group was collected through a questionnaire to evaluate comfort, usability, and satisfaction. The trial was prospectively registered at ClinicalTrials.gov (NCT06244719).

**Results:** The vibrotactile stimulation was safe and well-tolerated by stroke patients. No statistically significant differences were noted in FMA-UE motor scores and total MAS scores between the groups; however, exploratory analyses revealed a significant improvement in FMA-UE joint pain scores in the treatment group. A trend towards reduced wrist spasticity was observed in the treatment group, but this effect did not remain significant after correcting for multiple comparisons. Responder analyses showed a greater proportion of responders in the treatment group for both FMA-UE and MAS outcomes.

**Conclusions:** Our results show promise for a larger sample size study and follow-up work with longer durations of vibrotactile stimulation with gloves and shirts for upper extremity stroke rehabilitation. Although no significant improvements were observed in FMA-UE motor function or total MAS scores, vibrotactile stimulation was associated with reduced joint pain, potential benefits for distal spastic hypertonia, and increased responder rates.

## Introduction

Approximately 17 million people globally experience a first-ever stroke each year (Feigin et al., 2014). As many as 80% of stroke survivors have upper extremity impairments such as spasticity, weakness, abnormal muscle activity, limited tactile perception, and range of motion (Hayward et al., 2021). Six months post-stroke, some dexterity in the paretic upper extremity was found in 38% of patients, while only 11.6% of patients experienced a complete functional recovery (Kwakkel et al., 2015). Impairment of the upper extremity is often chronic and a cause of long-term disability (Ward, 2017). Most stroke survivors have difficulties using their impaired upper extremity in activities of daily living such as eating with utensils, dressing up, and opening a door. They view the recovery of the upper extremity as a priority to improve their quality of life (Au-Yeung and Hui-Chan, 2009). However, previous studies showed that in our current rehabilitation model, patients spend a limited time in therapy doing rehabilitation exercises (Lang et al., 2009), as our current rehabilitation model is primarily clinic-based and resource-intensive. This contrasts with the evidence that shows that high-intensity rehabilitation, early after stroke and over a continued period, is critical to maximize recovery (Hayward et al., 2021). Wearable technology has the potential to reduce this limitation of our current rehabilitation model.

Studies with magnetoencephalography indicate that beta-band responses to tactile fingers stimulation modulate sensorimotor excitability and are correlated with the recovery of hand dexterity and increased sensory map size in stroke patients (Laaksonen et al., 2012; Roiha et al., 2011). Vibratory stimuli applied to spastic upper extremity muscles for five minutes brought a significant reduction in muscle tonus, which lasted for at least 30 minutes after the stimulation ended (Noma et al., 2012). Transcranial magnetic stimulation after vibratory stimuli was applied for 10 minutes (3 times/day for 3 consecutive days) revealed a reduction in resting motor threshold and an increase in the motor map areas of the vibrated muscles that were correlated with a reduction in spasticity and an increase in motor function (Marconi et al., 2011). It was hypothesized that the improvements associated with vibrotactile stimulation were partially due to Ia afferent input that induced a functional restoration of inactivated, but preserved motor pathways and a rearrangement of motor cortical maps. A possible mechanism for the vibrotactile stimulation to reduce spasticity and prevent excessive agonist/antagonist co-contraction is that it induces an upregulation and downregulation of GABAergic circuits when controlling the pairs of antagonists. The benefits of treatment were observed in acute stroke patients when the stimulation was applied on two upper extremity muscles, specifically the flexor carpi radialis and biceps brachii (Toscano et al., 2019). Stimulating other muscles on the upper extremity was not investigated.

Spasticity affects 20-45% of chronic stroke survivors (Seim et al., 2022) and can impair hand use, hygiene, and comfort (Patel et al., 2019). The standard clinical treatment is intramuscular injection of botulinum toxin type A, which often yields limited results and causes weakness (Comella et al., 2021). Seim and colleagues conducted a clinical feasibility study with a vibrotactile stimulation glove and 16 one-year post-stroke patients (Seim et al., 2021). Stimulating the knuckles yielded significant reductions in spasticity, an increase in tactile perception, and upper extremity range of motion. In a follow-up study, it was found that vibrotactile stimulation yielded equal or more effective outcomes in spasticity than botulinum toxin (Seim et al., 2022). Stimulating other parts of the upper extremity or other locations on the hand, such as the fingertips, which have a large cortical representation (Penfield and Boldrey, 1937), has not yet been investigated with a wearable device, which could be used for at-home rehabilitation.

In this study, we investigated the effects of vibrotactile stimulation in the upper extremity with wearable devices (shirts and gloves). Vibrotactile stimulation has been shown to improve upper extremity function and spasticity (e.g., Marconi et al., 2011; Noma et al., 2009), but it has been largely limited to laboratory and clinical studies. In most prior studies, vibrotactile stimulation was only applied for a few minutes or days. The potential benefits of applying the stimulation for longer durations are not known. Wearable devices have the potential to increase the duration and length of the therapy, which seems critical to improve recovery (Hayward et al., 2021). Furthermore, given that post-stroke spasticity is common in the muscles of the upper and lower arms (Thibaut et al., 2013), our devices have the potential to yield further improvements than a simulation limited to the knuckles (Seim et al., 2021). The cost-effectiveness of the wearable devices (approximately $80) could allow the widespread adoption of the therapy.

We conducted a randomized controlled trial to assess the safety, feasibility, and efficacy of vibrotactile stimulation with wearable devices. Patients were recruited within 2 weeks post-stroke in the rehabilitation unit. The primary outcome measure was the FMA-UE motor score conducted at baseline and at discharge from the inpatient rehabilitation unit. Our secondary outcome measures included the MAS at the same two time points. A verbal survey was conducted to assess device tolerability. We hypothesized that the addition of vibrotactile stimulation to conventional rehabilitation therapy would result in greater improvements in upper-extremity motor recovery, as measured by the FMA-UE motor score. We further hypothesized that participants receiving vibrotactile stimulation would exhibit greater reductions in spastic hypertonia, as measured by the MAS.

### Study design

Between May 2024 and February 2026, we conducted a randomized controlled trial to test the superiority of vibrotactile stimulation in addition to conventional rehabilitation therapy to conventional rehabilitation therapy alone. A total of 63 stroke patients were assessed for eligibility, of whom 39 were excluded (n=29 did not meet the inclusion criteria and n=10 declined to participate; see Fig 1). Twenty-four participants were randomized 1:1 to a treatment group (vibrotactile stimulation in addition to conventional rehabilitation therapy) and a control group (conventional rehabilitation therapy alone). Block randomization (block size of 6) generated by a computer program was used to avoid bias in treatment allocation. Randomization was stratified according to baseline FMA-UE motor score (0–20, 21–40, and 41-58). The random allocation sequence was computer-generated by a research assistant using block randomization. The same research assistant enrolled participants and assigned them to the intervention groups. Allocation concealment was not implemented because the individual responsible for generating the random allocation sequence also performed participant enrolment and assignment. Nevertheless, participants were enrolled sequentially as they were admitted to the rehabilitation unit using the predefined eligibility criteria, thereby reducing the risk of selection bias. Our study workflow is illustrated in Fig 2. In the treatment group, stroke patients wore the glove on their impaired hand for 45 minutes and the shirt for another 45 minutes—in no specific order—5 days a week during their stay at the rehabilitation unit. Patients wore the devices after they had finished their daily 3 hours of conventional rehabilitation therapy (i.e., standard of care) at 3 pm. The standard of care consists of an average of 3 hours of combined physical therapy, occupational therapy, and speech therapy per day, 7 days per week. Our objective was to have the therapy delivered early after stroke, as animal and now human studies indicate that intense rehabilitation during a critical time window after stroke promotes greater recovery (Ballester et al., 2022; Patel et al., 2019). We wanted the study to occur at the rehabilitation unit so that any adverse event could be addressed quickly. Our devices included electronic components, and there was a minimal risk of electric shock or other unanticipated complications.

**Fig 1.**
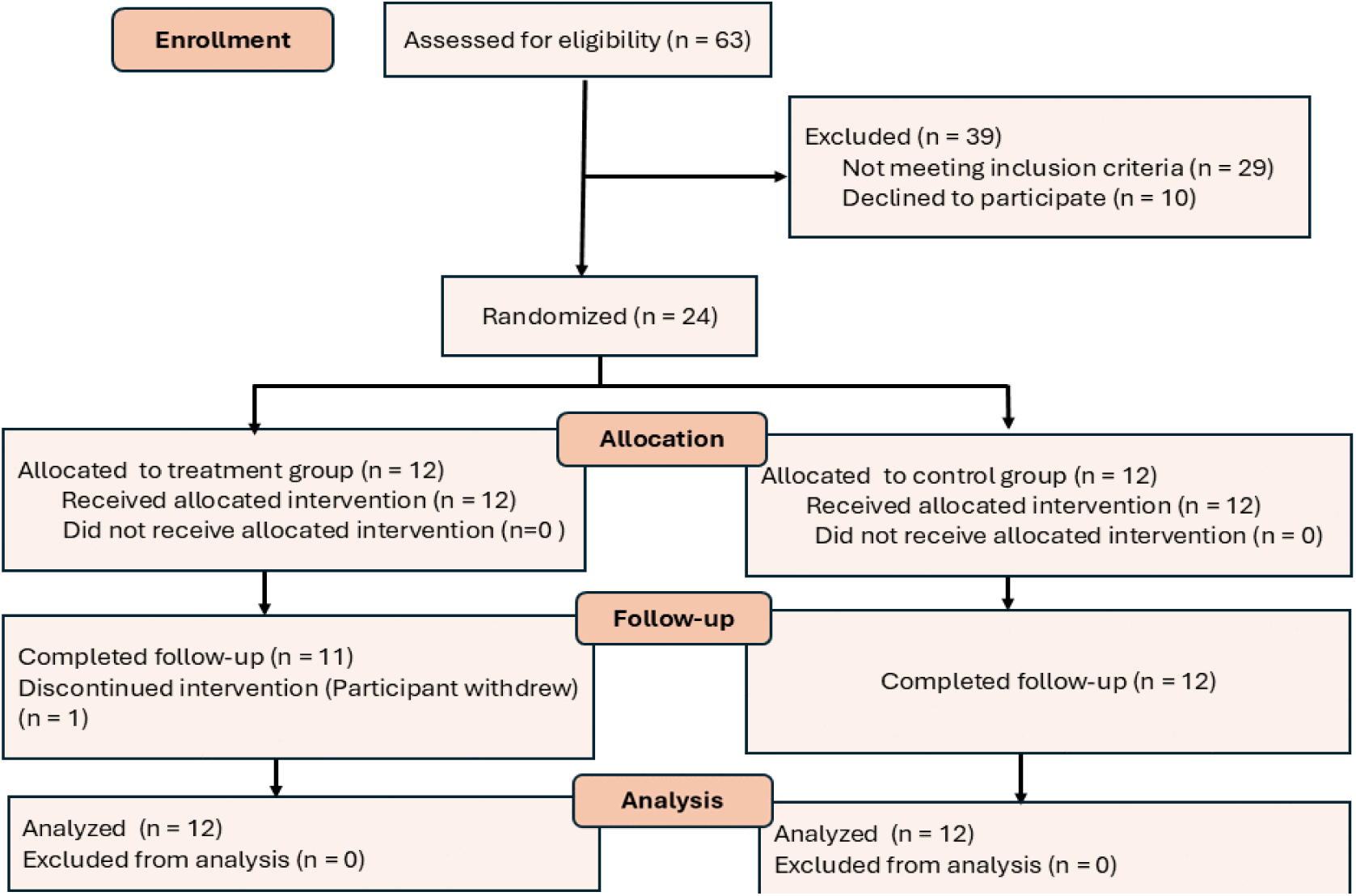
Participant flow through the study. A total of 63 stroke patients were assessed for eligibility. Of these, 39 were excluded as 29 did not meet the inclusion criteria and 10 declined to participate. The remaining 24 participants were randomized in a 1:1 ratio to the treatment group (n = 12) or control group (n = 12). One participant in the treatment group withdrew before completing the treatment. All 24 randomized participants were included in the final analysis.

**Fig 2.**
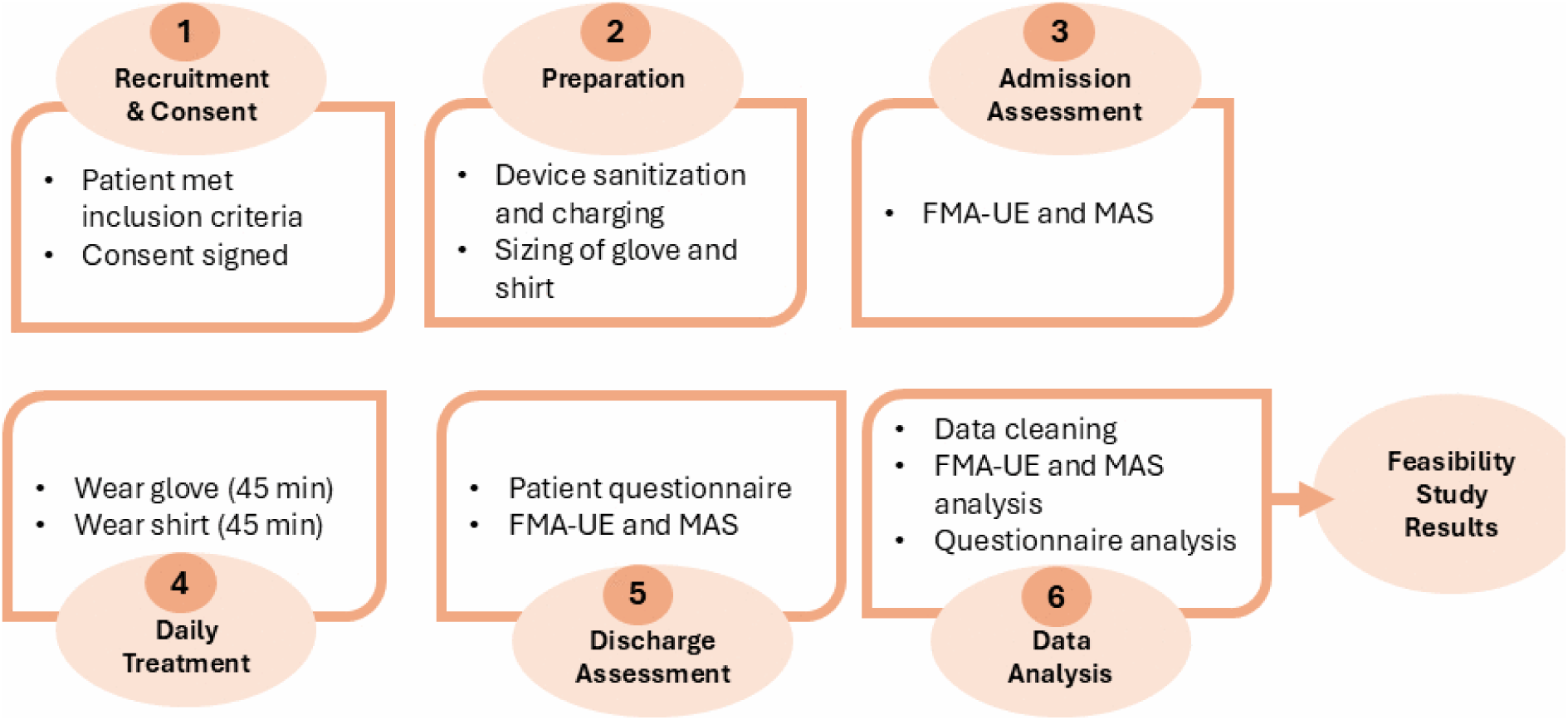
Overview of study workflow. The study workflow included six phases: (1) recruitment and consent form signature for eligible participants, (2) preparation of the devices including sanitization, charging, and sizing, (3) FMA-UE and MAS admission assessments, (4) daily treatment involving 45 minutes of glove use and 45 minutes of shirt use following conventional rehabilitation therapy, (5) FMA-UE and MAS discharge assessments, and (6) data analysis, including feasibility, clinical outcome, and questionnaire analyses.

The primary outcome measure was the FMA-UE conducted at baseline and at discharge from the inpatient rehabilitation unit. Our secondary outcome measures included the MAS, the number of participants with an adverse event, and the number of participants who completed and tolerated the vibrotactile stimulation treatment. The FMA-UE and MAS were conducted by OTs who were not involved with the study. The participants did not wear the vibrotactile stimulation devices when the assessments were conducted. Assessor blinding could not be ensured because the OTs may have observed the participants wearing the vibrotactile stimulation devices on days preceding the assessments. We also included the FMA-UE sensation, passive joint motion, and joint pain scores, and the MAS for the fingers, wrist, and elbow scores as exploratory outcome measures. A verbal survey conducted at discharge assessed how patients tolerated the therapy and was used to get feedback on device comfort and usability. Therapy compliance was assessed by the survey and feedback from our research assistants. The time points were chosen to capture critical information for each measure while minimizing the participant testing burden.

Our vibrotactile wearable devices were presented to stroke participants at the inpatient rehabilitation unit who fulfilled the inclusion criteria. Participants who agreed to participate in the study signed a consent form personally. Some participants who suffered from cognitive impairments after stroke consented with the help of their legally authorized representative, after verbally ensuring that they had understood and agreed to participate in the study. All devices were sanitized with an alcohol-based cleaner before being provided to participants. In addition, devices were hand-washed every three days (or more often if needed), and the vibrotactile motors were sanitized with an alcohol-based cleaner. The participants were allowed to wear a thin shirt below the vibrotactile stimulation shirt. The devices were charged every night as they have rechargeable batteries. On the first study visit, we chose the proper size of glove and shirt to be assigned to each participant according to their limb size. Our research assistants were present with the patients when wearing the devices to ensure no adverse event occurred, help them wear and remove the devices (e.g., when eating or going to the restroom), charge the batteries nightly, and clean the devices.

We have chosen the sample size of our study based on a power analysis to detect the minimally clinically important difference of 7 in the FMA-UE score between the vibrotactile stimulation therapy and the conventional rehabilitation therapy (Krakauer et al., 2021). Ten participants per group provided 84% power (effect SD at 5) based on a 2-sample t-test with a 2-sided α level at 0.05. Our study was conducted with 12 participants per group to account for participant attrition. The study was reviewed and approved by Colorado Multiple Institutional Review Board (COMIRB Protocol 23-0783) and registered under ClinicalTrials.org (Identifier: NCT06244719; https://clinicaltrials.gov/study/NCT06244719) on January 29, 2024, prior to enrolling the first participant.

### Participants

This pilot study was performed in Denver, Colorado. A total of 24 ischemic (hemispheric or brainstem, detected by magnetic resonance imaging or computer tomography) stroke patients with upper extremity deficits (ages 45-85 years old) were included. All participants were recruited from the UCHealth rehabilitation units at the University of Colorado Broomfield Hospital and University of Colorado Hospital, Anschutz Medical Campus. Participants at both locations undergo the same conventional rehabilitation therapy program. All participants provided written consent to participate in the study. In the treatment group, after participants completed their conventional rehabilitation therapy for the day, they wore a vibrotactile glove and shirt for 45 minutes each (not wearing the devices at the same time), five days a week. Participants were permitted to take a break in between wearing the devices. The duration of our vibrotactile stimulation treatment was 3 hours and 45 minutes per week for each device, totalling 7 hours and 30 minutes of stimulation per week. In the control group, participants proceeded with their conventional rehabilitation therapy. The vibrotactile stimulation devices were not used with the participants in this group.

### Selection criteria

Study inclusion criteria included being between 45 and 85 years old, having a unilateral left or right-sided ischemic stroke within the past 2 weeks, having a FMA-UE motor score between 0 and 58, and expected to stay at least 1 week in the inpatient rehabilitation unit. Exclusion criteria included patients who are involved in other clinical trials or undergoing anti-spasticity therapy during the study period, patients with communicable diseases or "yellow gown isolation" such as Clostridium Difficile and Methicillin-resistant Staphylococcus aureu, patients dependent on pacemakers or defibrillators, or have lymphedema or an arteriovenous fistula for dialysis in one arm.

### Wearable devices

Our shirts and gloves were designed to provide vibrotactile stimulation to stroke patients during their stay at the inpatient rehabilitation unit. The vibration motors were coin-type eccentric rotating mass (ERM) motors (model HD-EMC1003-LW40-R, PUI Audio Inc.), rated at a voltage of 3.0 V DC (operating voltage range: 2.5 V DC to 3.5 V DC). The motors had a rated speed of 12,000 ± 3,000 RPM and a maximum current of 85 mA at 3.0 V DC. Our gloves and shirts had 5 motors to stimulate the fingertips and 18 motors to stimulate the impaired upper extremity.

### Glove design

A fabric wearable glove was powered with a LilyPad Arduino (SparkFun, Product ID 13342, 2.7 V – 5.5 V) microcontroller and a rechargeable 3.7 V, 1000 mAh lithium-iron battery (SparkFun, Product ID 13813) connected via a LilyPad power supply board (SparkFun, Product ID 11893). The motors had a rated speed of 12,000 ± 3,000 RPM. 3D printed motor casings were used to hold the motors and were formed to fit each fingertip with heat-softening and sanding for comfort. The battery was covered with a custom 3D printed case and fastened using fabric Velcro. All electronics were covered with non-conductive foam padding for electrical insulation and comfort. Our vibrotactile stimulation gloves were built for three sizes: large, medium, and small. The glove design is illustrated in Figs 3A and 3B. Gloves were designed to be portable, lightweight (100 g), and washable. Our devices stimulate the fingertips because of the large cortical areas that are dedicated to their sensory processing (Härtner et al., 2021; Janko et al., 2022). The fingertips were stimulated randomly, one fingertip at a time.

**Fig 3.**
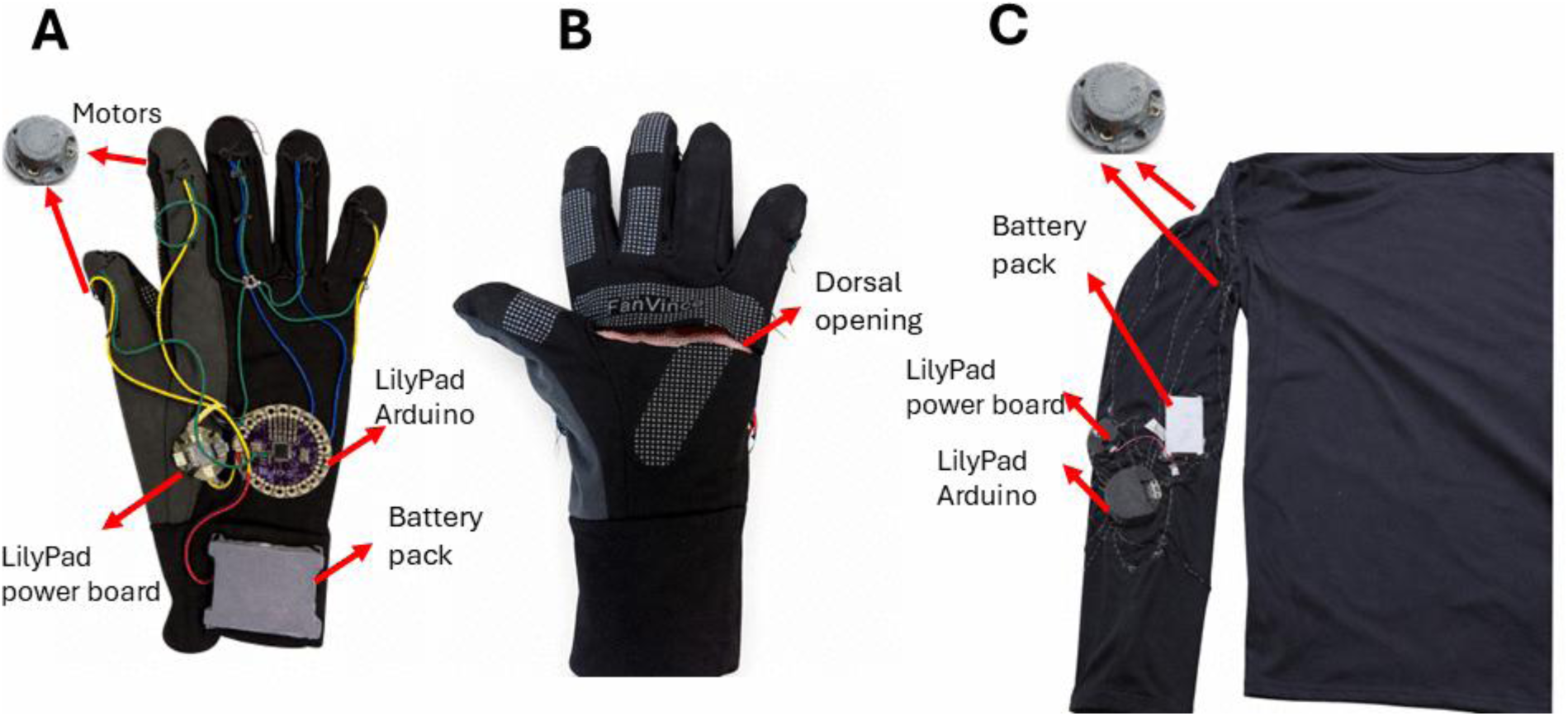
Design of the vibrotactile glove and shirt devices. (A) Front side of the vibrotactile stimulation glove showing the main hardware components, including the vibration motors, LilyPad Arduino, LilyPad power board, and battery pack, (B) back side of the glove showing the opening along the dorsal surface, which was incorporated to facilitate donning and doffing by stroke patients, (C) vibrotactile stimulation shirt (medium size) showing the integrated electronic components, including the vibration motors, LilyPad Arduino, LilyPad power board, and battery pack.

### Shirt design

Shirts were built for four different sizes: x-large, large, medium, small, and x-small. All the components were placed either on the left or right sleeve to stimulate the patient’s affected limb. The same electronic components for the gloves were used to build the shirts (microcontroller, battery, etc.). The 18 motors were placed around the sleeve: six on the front forearm, two on the back forearm, four on the front upper arm, three on the back upper arm, and three on the shoulder. Fig 3C depicts the designed shirt.

### Motor vibration frequency

In the first stages of our system design, the vibration motors were controlled with the default Arduino function for pulse width modulation (PWM). The motor vibration intensity was variable throughout the session due to the dynamic control of the PWM period, resulting in the motors receiving uneven power. The unstable vibration intensity was corrected by implementing a PWM-based vibration delivery system. A timer interrupt function was implemented for controlling the vibration motors by generating a fixed PWM signal. The motors were driven at an operating voltage of approximately 3.0 V DC (within a 2.5–3.5 V range). To achieve a motor vibration frequency of 80 to 100 Hz, a 35% duty cycle was used. This was determined by attaching an accelerometer to the vibration motors and computing the vibration frequency with the Fourier Transform.

### Arduino implementation

We built a customized Arduino-based control system to provide the vibrotactile stimulation through our wearable devices. Our devices are designed to activate one motor at a time with the proper intensity, using a random vibration pattern. Each selected motor remained active for 1 s before being deactivated and replaced by another randomly selected motor. As shown in Fig 4, the Arduino code operates our vibrotactile devices (gloves and shirts), initializes the motor pins, and sets vibration frequency via the duty cycle. A timer interrupt produces the PWM signals to alternate the motors that are on and off, enabling vibration delivery without loop blocking.

**Fig 4.**
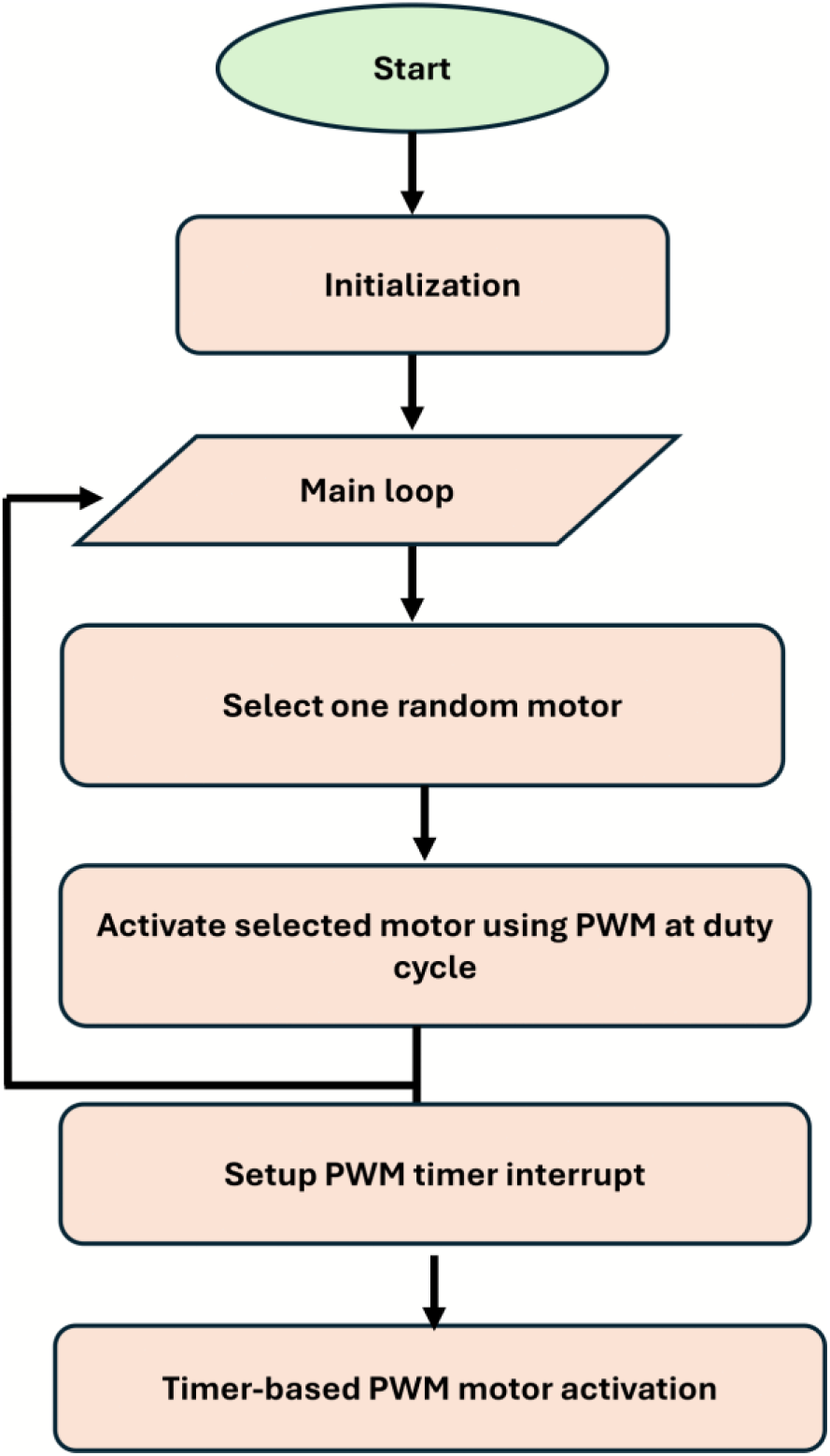
Flowchart of the vibrotactile stimulation control algorithm used in the glove and shirt devices. Following initialization, the system configures the number of available vibration motors (5 for the glove and 18 for the shirt) and assigns the stimulation parameters. During the main control loop, a motor is randomly selected and activated using PWM at the specified duty cycle. A timer interrupt is then configured to control motor activation and deactivation independently of the main program loop.

### Data analysis

Descriptive statistics were employed to describe the demographic and clinical parameters at baseline, such as sex, age, stroke diagnosis, and length of stay in the rehabilitation unit for both groups. Data from all participants were analyzed, including from one participant who withdrew from the treatment group before discharge from the rehabilitation unit. For this participant who dropped out, the last available values for the FMA-UE and MAS were carried forward. The Mann–Whitney U test was used to ensure that the baseline motor scores are balanced between groups at the start. All our statistical analyses were implemented in Python (version 3.11.3).

The FMA-UE and MAS scores were analyzed to compute upper extremity motor impairment and spastic hypertonia at admission and discharge. A change score was calculated for each participant (post - baseline). Positive and negative values indicated improvement on the FMA-UE and MAS, respectively. Changes within-group were assessed with the Wilcoxon signed-rank test to determine whether the median change differed from zero. Between-group differences (treatment vs control) in the change scores were analyzed using the Mann–Whitney U test (MacFarland and Yates, 2016). In this manuscript, effect sizes were reported with the Hodges–Lehmann (HL) estimate (treatment – control for between-group and post – baseline for within-group comparisons), where positive and negative values indicate greater improvement in the treatment group for FMA-UE and MAS, respectively. Standardized effect sizes were calculated as *r* = |*z*|/√*n*. For the MAS, flexor and extensor ratings were averaged for each joint to obtain a single score at baseline and discharge. For the exploratory outcome measures, p-values were adjusted for multiple comparisons with the Benjamini–Hochberg false discovery rate (FDR) procedure. Both raw and adjusted p-values are reported in this study. We considered a p-value less than 0.05 to be statistically significant.

Responder analyses were performed to quantify clinically meaningful improvements. For the FMA-UE motor domain, responders were defined as participants who improved by at least 5 points, based on published minimal clinically important difference estimates (Page et al., 2012). For the sensation, passive joint motion, and joint pain domains, responders were defined as participants who improved by at least 1 point (Fugl-Meyer et al., 1975). For the MAS, responders were defined as participants who demonstrated a reduction of at least 1 point, exceeding published minimal clinically important difference estimates for stroke populations (Chen et al., 2019). Responder proportions were compared between groups using Fisher’s exact test.

### Participant feedback questionnaire

At discharge, each participant in the treatment group completed a verbal questionnaire to evaluate the feasibility and usability of our devices. The questionnaire addressed the following aspects: device comfort, tolerability, wearability, sensation of vibration, freedom of movement, and general user experience. Further, participants were encouraged to provide any feedback and suggestions to improve our devices.

## Results

### Participant characteristics

Table 1 illustrates the demographic and clinical characteristics of participants. Participants were recruited from the Anschutz Medical Campus (treatment n=8; control n=7) and Broomfield Hospital (treatment n=4; control n =5). Study participants were recruited on average 10.21 ± 7.62 days post-stroke and had an average length of stay of 15.25 ± 4.46 days at the rehabilitation unit. Participants in the treatment group received vibrotactile stimulation for an average of 6.3 ± 1.0 days during their stay in the rehabilitation unit. To check whether baseline (i.e., at admission) motor scores were balanced between groups, the Mann–Whitney U test was used. The baseline FMA-UE motor scores of the two groups were not significantly different from each other (Mann–Whitney U = 67.50, p = 0.814, r = 0.05, HL estimate = 0.0 points, 95% CI −10.0 to 6.0, see Fig. 5). The median values were 7.5 for the treatment group and 7.0 for the control group.

**Fig 5.**
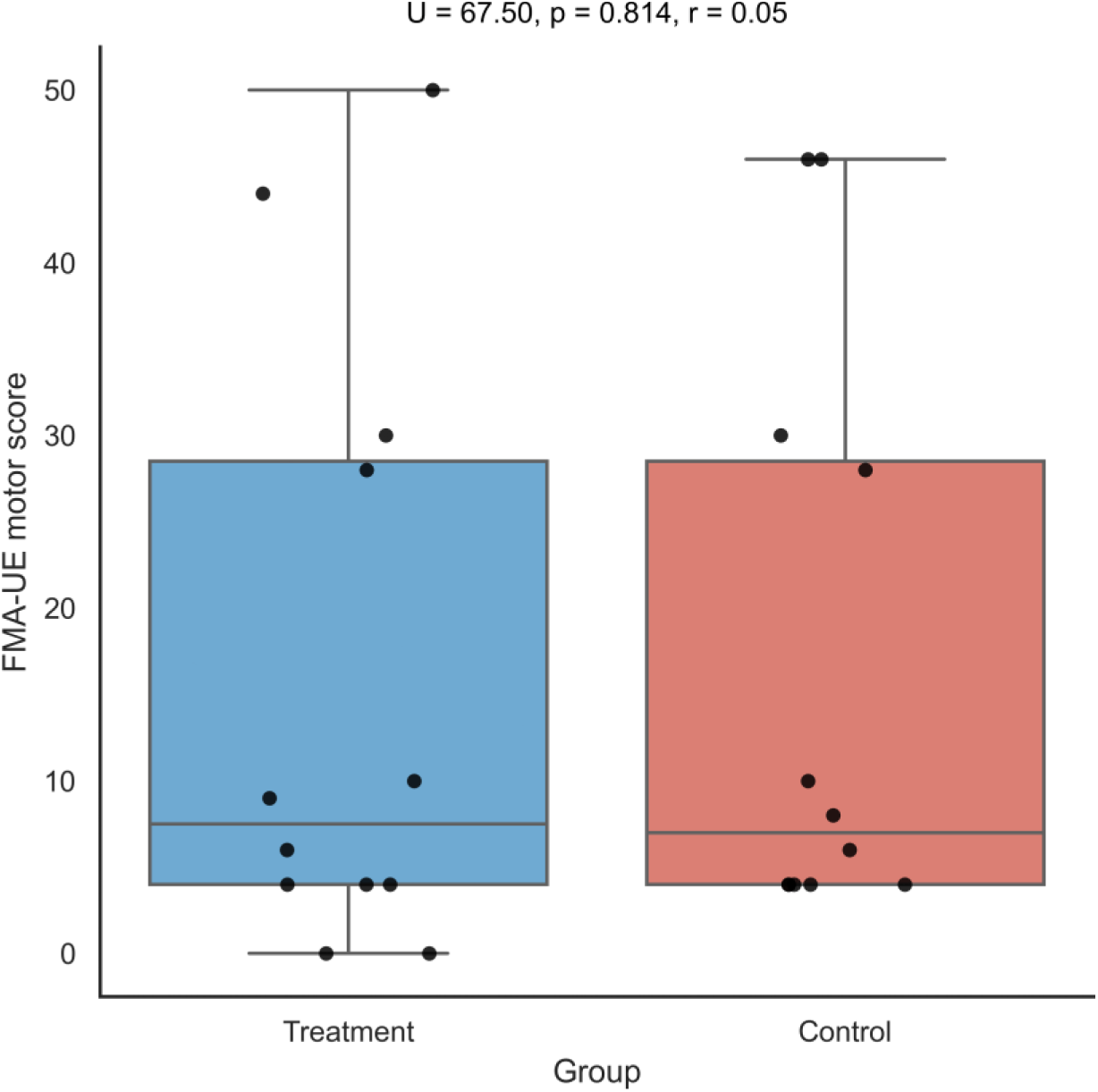
Baseline equivalence between treatment and control groups. Boxplots show baseline FMA-UE motor scores at admission for participants assigned to the treatment group (n = 12) and control group (n = 12). Dots represent individual participants, and the horizontal line within each box indicates the median score. No significant difference was observed between groups at baseline (Mann–Whitney U = 67.50, p = 0.814, r = 0.05). Effect sizes are reported as r.

**Table 1.**
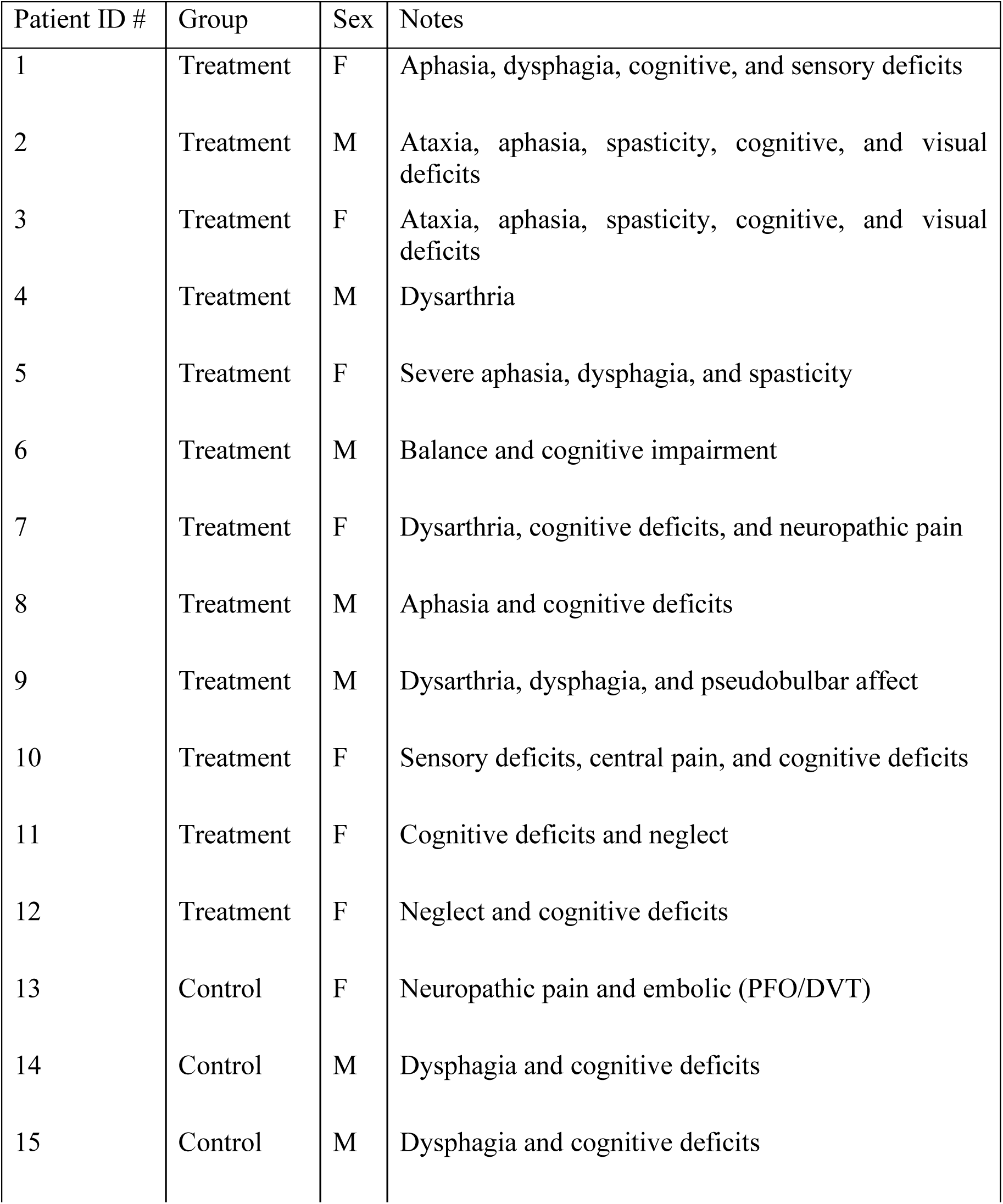
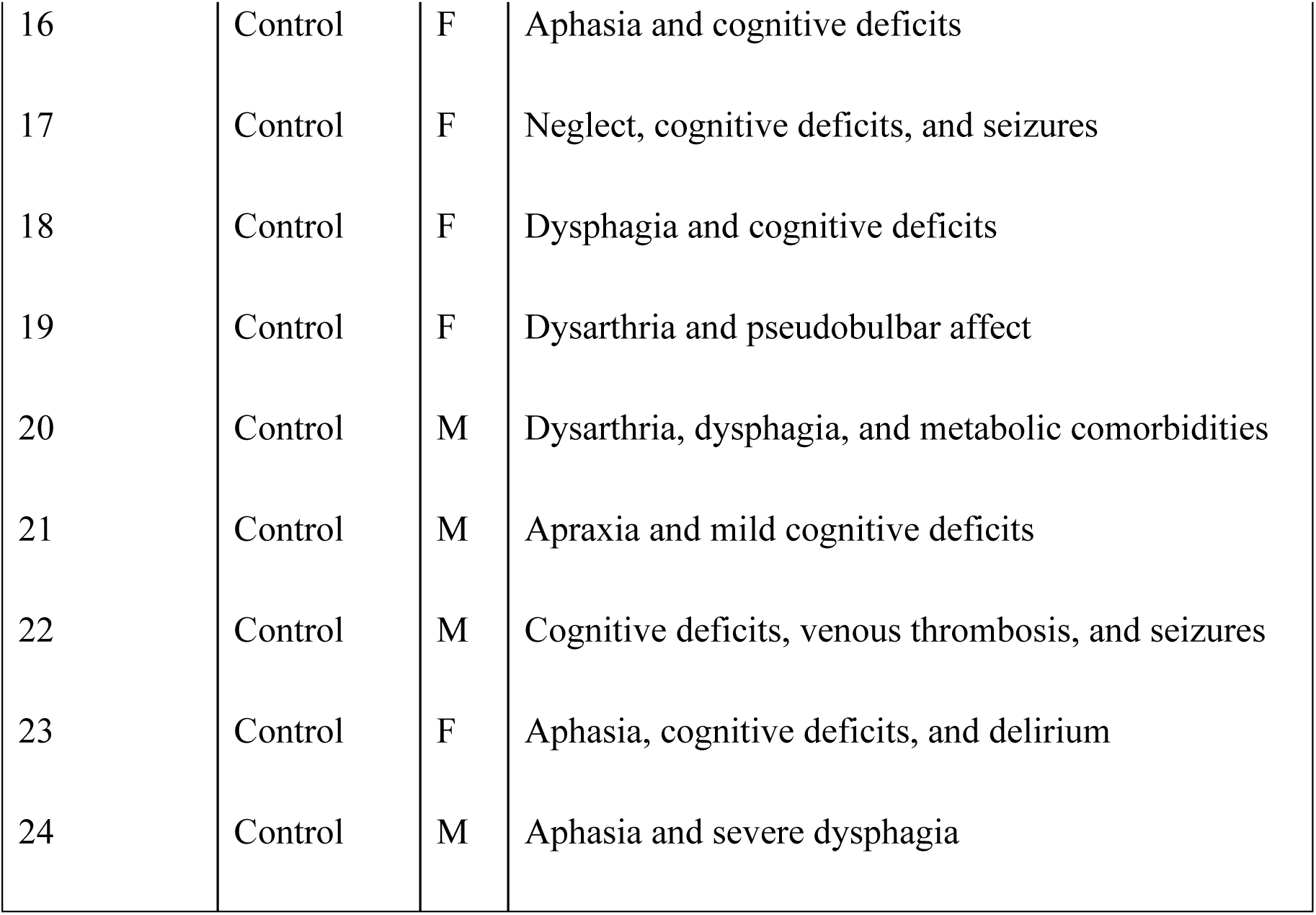
Demographics and participant characteristics at baseline.

| Patient ID # | Group | Sex | Notes |
| --- | --- | --- | --- |
| 1 | Treatment | F | Aphasia, dysphagia, cognitive, and sensory deficits |
| 2 | Treatment | M | Ataxia, aphasia, spasticity, cognitive, and visual deficits |
| 3 | Treatment | F | Ataxia, aphasia, spasticity, cognitive, and visual deficits |
| 4 | Treatment | M | Dysarthria |
| 5 | Treatment | F | Severe aphasia, dysphagia, and spasticity |
| 6 | Treatment | M | Balance and cognitive impairment |
| 7 | Treatment | F | Dysarthria, cognitive deficits, and neuropathic pain |
| 8 | Treatment | M | Aphasia and cognitive deficits |
| 9 | Treatment | M | Dysarthria, dysphagia, and pseudobulbar affect |
| 10 | Treatment | F | Sensory deficits, central pain, and cognitive deficits |
| 11 | Treatment | F | Cognitive deficits and neglect |
| 12 | Treatment | F | Neglect and cognitive deficits |
| 13 | Control | F | Neuropathic pain and embolic (PFO/DVT) |
| 14 | Control | M | Dysphagia and cognitive deficits |
| 15 | Control | M | Dysphagia and cognitive deficits |
| 16 | Control | F | Aphasia and cognitive deficits |
| 17 | Control | F | Neglect, cognitive deficits, and seizures |
| 18 | Control | F | Dysphagia and cognitive deficits |
| 19 | Control | F | Dysarthria and pseudobulbar affect |
| 20 | Control | M | Dysarthria, dysphagia, and metabolic comorbidities |
| 21 | Control | M | Apraxia and mild cognitive deficits |
| 22 | Control | M | Cognitive deficits, venous thrombosis, and seizures |
| 23 | Control | F | Aphasia, cognitive deficits, and delirium |
| 24 | Control | M | Aphasia and severe dysphagia |

### Within-group improvement

Changes from baseline to post within each group across the four FMA-UE domains (motor function, sensation, passive joint motion, and joint pain) were analyzed (see Fig 6). For motor function (see Fig 6A), both groups showed significant improvements (<u>treatment</u>: p = 0.003, r = 0.85; HL estimate = 9.25 points, 95% CI 3.25 to 20.50; <u>control</u>: p = 0.002, r = 0.88, HL estimate = 6.25 points, 95% CI 3.25 to 20.50). The treatment group showed nominal improvements in sensation (p = 0.094, p_adj_ = 0.094, r = 0.84, HL estimate = 1.5 points, 95% CI 0.00 to3.00, see Fig 6B), passive joint motion (p = 0.070, p_adj_ = 0.094, r = 0.78, HL estimate = 1.00 point, 95% CI 0.00 to 2.50, see Fig 6C) and joint pain (p = 0.031, p_adj_ = 0.094, r = 0.78, HL estimate = 2.00 points, 95% CI 0.25 to 4.50, see Fig 6D), but without reaching significance after FDR-correction. In contrast, the control group did not approach significance (<u>sensation</u>: p = 0.812, p_adj_ = 0.812, r = 0.69, HL estimate = 0.00 points, 95% CI −0.50 to 1.00, see Fig. 6B; <u>passive joint motion</u>: p = 0.375, p_adj_ = 0.562, r = 0.77, HL estimate = 0.00 points, 95% CI 0.00 to 1.00, see Fig 6C; <u>joint pain</u>: p = 0.375, p_adj_ = 0.562, r = 0.85, HL estimate = 0.00 points, 95% CI −1.00 to 0.00, see Fig 6D).

**Fig 6.**
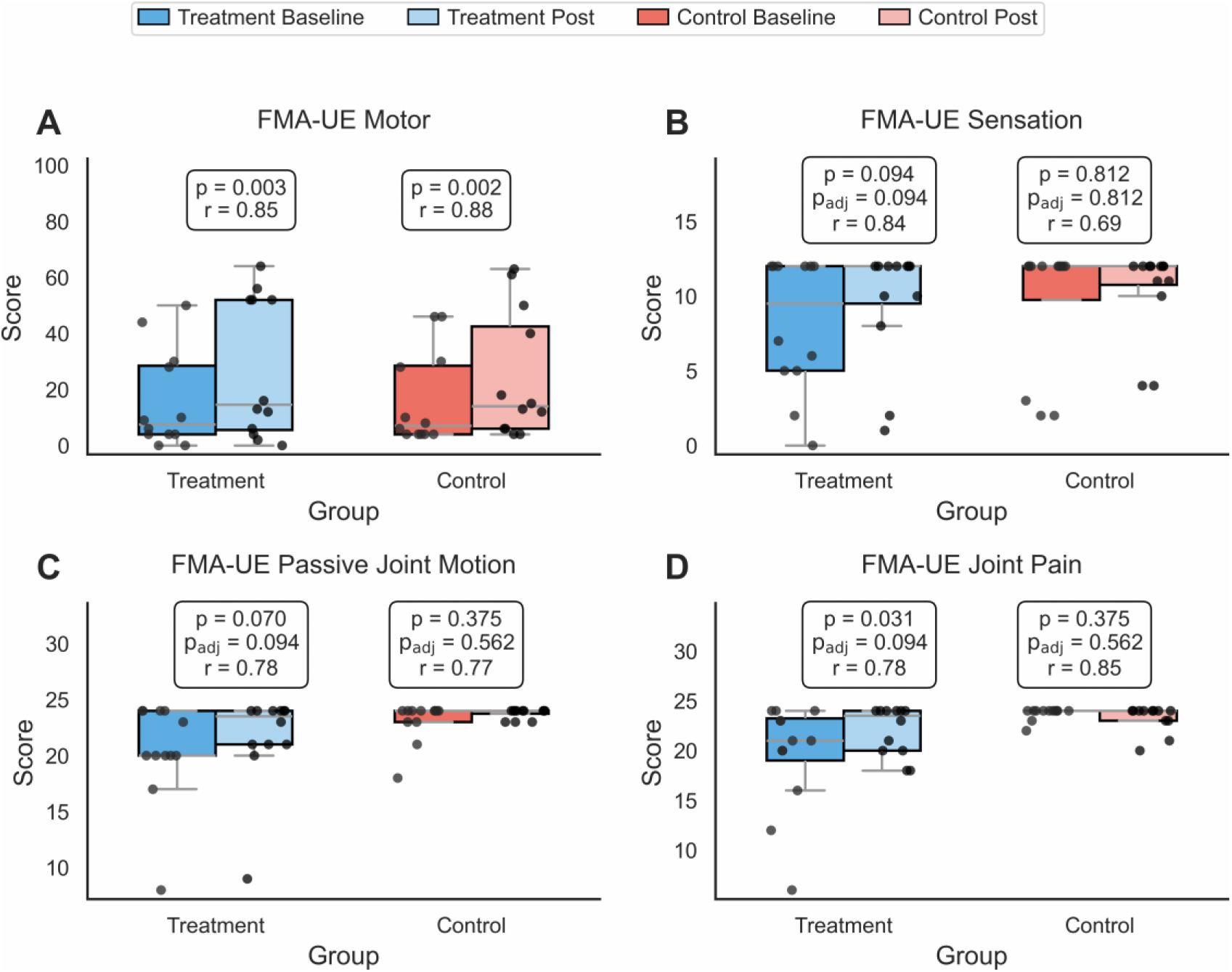
FMA-UE improvement scores within groups from admission (baseline) to discharge (post). (A) motor function, (B) sensation, (C) passive joint motion, and (D) joint pain. Boxplots show the distribution of scores at baseline and discharge; horizontal lines indicate the median, and dots represent individual participants. Within-group changes were evaluated using the Wilcoxon signed-rank test. For the primary outcome (motor function), the raw p-value (p) is reported. For the exploratory outcomes (sensation, passive joint motion, and joint pain), both raw and FDR-adjusted (p_adj_) p-values are reported. Effect sizes are reported as r. Significant improvements in motor function were observed in both the treatment (p = 0.003, r = 0.85) and control (p = 0.002, r = 0.88) groups.

### Between group improvement

We then compared changes between groups across all FMA-UE domains (see Fig 7). No significant between-group differences were found for motor function (U = 81.50, p = 0.602, r = 0.11, HL estimate = 2.0 points, 95% CI −4.0 to 11.0), sensation (U = 90.00, p = 0.280, p_adj_ = 0.280, r = 0.21, HL estimate = 0.0 points, 95% CI 0.0 to 3.0), or passive joint motion (U = 91.00, p = 0.265, p_adj_ = 0.280, r = 0.22, HL estimate = 1.0 point, 95% CI -0.5 to 3.0). However, joint pain showed a significant difference between groups (U = 117.00, p = 0.007, p_adj_ = 0.020, r = 0.53, HL estimate = 3 points, 95% CI 0.0 to 4.0), with greater improvement in the treatment group. Additional participant-level FMA-UE changes are shown in S1 Fig.

**Fig 7.**
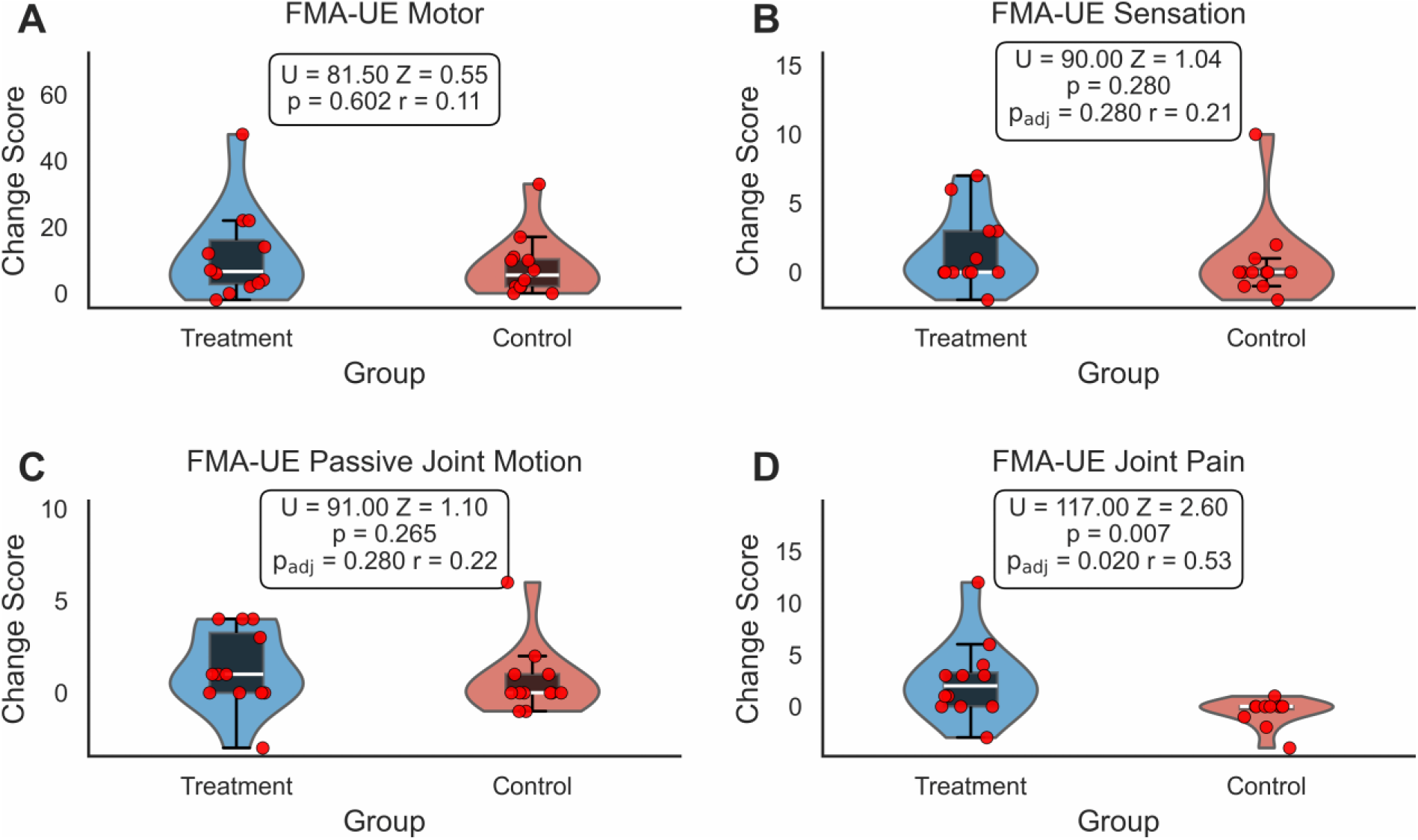
Changes in FMA-UE scores between groups from admission to discharge. Violin plots show the distribution of individual change scores, boxplots show the interquartile range, the white line indicates the median, and red dots represent individual participants. (A) Motor function, (B) sensation, (C) passive joint motion, and (D) joint pain. No significant between-group differences were observed for motor function (Mann–Whitney U = 81.50, p = 0.602, r = 0.11), sensation (U = 90.00, p = 0.280, p_adj_ = 0.280, r = 0.21), or passive joint motion (U = 91.00, p = 0.265, p_adj_ = 0.280, r = 0.22). A significant between-group difference was observed for joint pain (U = 117.00, p = 0.007, p_adj_ = 0.020, r = 0.53), with greater improvement in the treatment group.

### FMA-UE responder results

For the FMA-UE domains of motor function, sensation, and passive joint motion, responder rates were higher in the treatment group (58.3%, 41.7%, and 58.3%, respectively) compared to the control group (50.0%, 25.0%, and 33.3%), as shown in Fig 8. However, these differences were not statistically significant (<u>motor</u>: p = 1.000, HL estimate = 8.3%, 95% CI −27.8 to 41.6; <u>sensation</u>: p = 0.667, p_adj_ = 0.667, HL estimate = 16.7%, 95% CI −19.3 to 47.6; <u>passive joint motion</u>: p = 0.414, p_adj_ = 0.620, HL estimate = 25.0%, 95% CI −13.2 to 54.7). For joint pain, the responder rate was substantially higher in the treatment group (66.7%) than in the control group (8.3%), and this difference remained significant after FDR-adjustment (p = 0.009, p_adj_ = 0.028, HL estimate = 58.3%, 95% CI 19.7 to 79.0).

**Fig 8.**
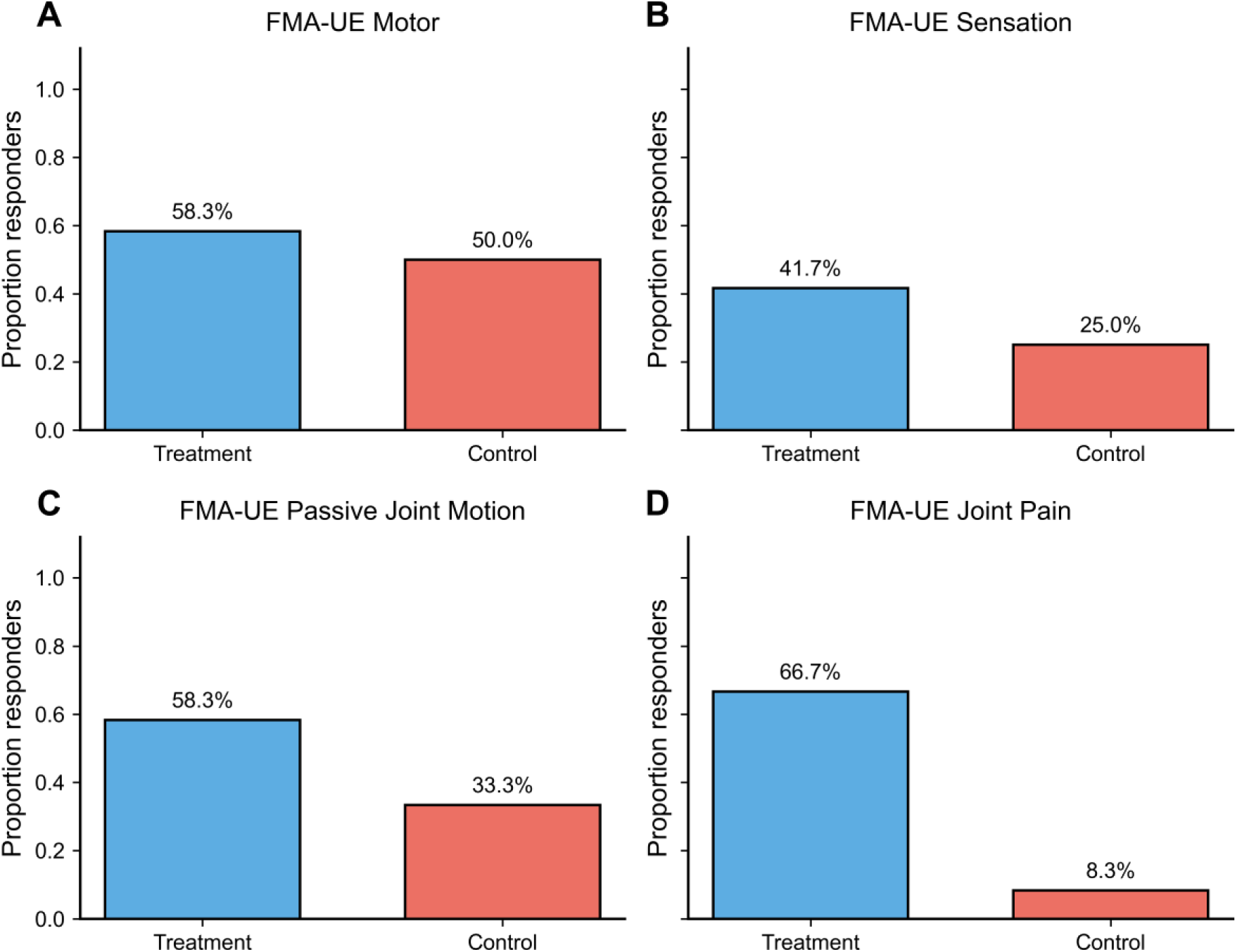
Responder analysis across FMA-UE domains. Bars represent the proportion of responders in the treatment and control groups. (A) Motor function: 58.3% of treatment participants and 50.0% of control participants were classified as responders (Fisher’s exact test, p = 1.000), (B) sensation: 41.7% of treatment participants and 25.0% of control participants were responders (p = 0.667, p_adj_ = 0.667), (C) passive joint motion: 58.3% of treatment participants and 33.3% of control participants were responders (p = 0.414, p_adj_ = 0.620), and (D) joint pain: 66.7% of treatment participants and 8.3% of control participants were responders (p = 0.009, p_adj_ = 0.028).

### MAS results

The analysis comparing total MAS change between groups showed no significant difference (p = 0.254, r = 0.24, HL estimate = −0.17 points, 95% CI −0.67 to 0.17, see Fig 9A). We then examined the changes in the individual MAS domains (fingers, wrist, and elbow). The treatment group had significantly improved MAS wrist scores compared to the control group (p = 0.040, r = 0.39; HL estimate = −0.50 points, 95% CI −1.0 to 0.0, see Fig 9C). However, this result did not remain significant after correcting for multiple comparisons (p_adj_ = 0.121). No significant between-group differences were observed in the scores for the fingers MAS (p = 0.449, p_adj_ = 0.673, r = 0.14, HL estimate = 0.0 points, 95% CI −0.75 to 0.0). No significant between-group difference was observed for elbow MAS (p = 0.878, p_adj_ = 0.878, r = 0.04, HL estimate = 0.0 points, 95% CI −0.50 to 0.50), as shown in Figs 9B and 9D. Additional participant-level MAS changes are shown in S2 Fig.

**Fig 9.**
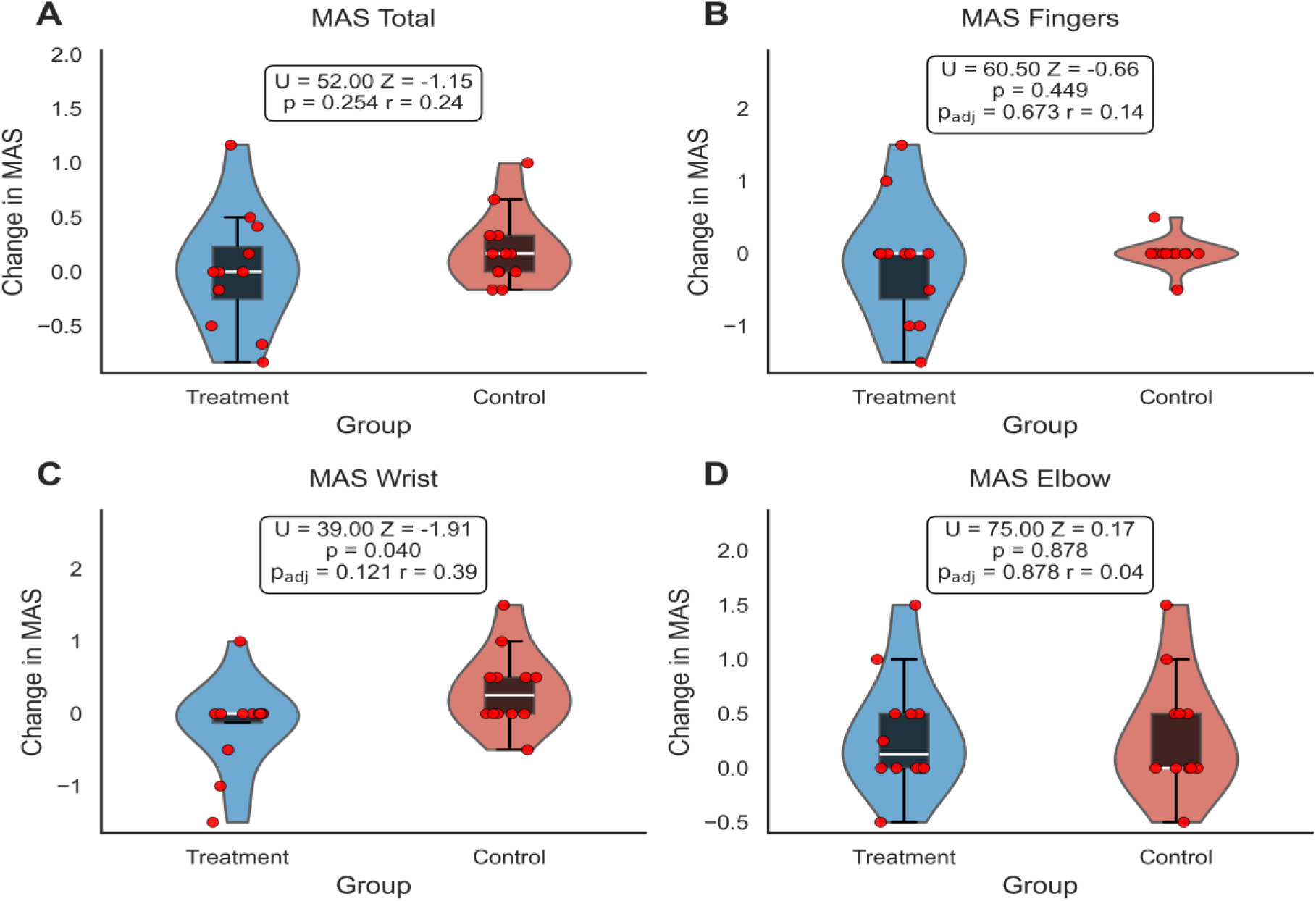
Changes in MAS scores between treatment and control groups from admission to discharge. Violin plots show the distribution of individual change scores, boxplots show the interquartile range, the white line indicates the median, and red dots represent individual participants. (A) Total MAS, (B) fingers, (C) wrist, and (D) elbow. No significant between-group differences were observed for total MAS (Mann–Whitney U = 52.00, p = 0.254, r = 0.24), fingers MAS (U = 60.50, p = 0.449, p_adj_ = 0.673, r = 0.14), or elbow MAS (U = 75.00, p = 0.878, p_adj_ = 0.878, r = 0.04). For the wrist MAS, the unadjusted analysis suggested a significant between-group difference with improvement in the treatment group (U = 39.00, p = 0.040, r = 0.39); however, the result did not retain significance after correcting for multiple comparisons (p_adj_ = 0.121).

### MAS responder results

We analyzed the proportion of responders, defined as participants with a ≥1-point reduction in MAS for the fingers, elbow, or wrist, in the control and treatment groups (see Fig 10). No responders were observed in the control group for any of the three regions examined. In the treatment group, responder rates were 25.0% (fingers), 16.7% (wrist), and 0.0% (elbow). However, the differences between groups were not statistically significant (<u>fingers</u>: p = 0.217, p_adj_ = 0.652, HL estimate = 25.0%, 95% CI −4.1 to 53.2; <u>wrist</u>: p = 0.478, p_adj_ = 0.717, HL estimate = 16.7%, 95% CI −10.4 to 44.8; <u>elbow</u>: p = 1.000, p_adj_ = 1.000, HL estimate=0.0%, 95% CI −24.2 to 24.2).

**Fig 10.**
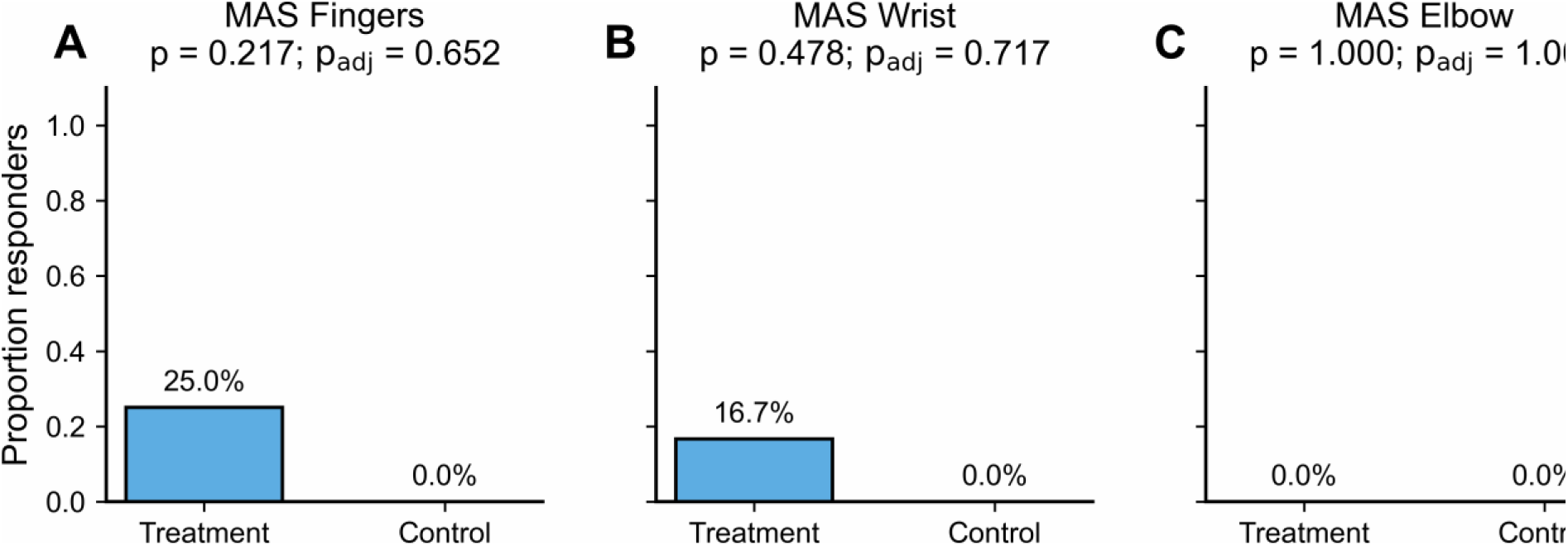
MAS responder analysis in the treatment and control groups. Bars represent the proportion of responders in each group. (A) Fingers: 25.0% of treatment participants and 0.0% of control participants were classified as responders (Fisher’s exact test, p = 0.217, padj = 0.652). (B) Wrist: 16.7% of treatment participants and 0.0% of control participants were responders (p = 0.478, padj = 0.652). (C) Elbow: no responders were observed in either group (p = 1.000, padj = 1.000).

### Questionnaire results

A majority of the participants in the treatment group reported the devices to be comfortable (10 out of 12 participants). Two participants reported uncomfortable sweating of the hands while wearing the gloves for 45 minutes. Moreover, one participant reported undesirable itching from the motors embedded in the shirts. The vibration itself was found to be noticeable and comfortable. All participants found improvement in their upper limbs’ sensation and decreased levels of tingling sensation from the vibrotactile stimulation treatment. All participants affirmed that they would use the devices in their own homes after discharge from the rehabilitation unit. One participant withdrew from the study after 4 days of vibrotactile stimulation, stating being too tired to receive 1.5 hours of stimulation after a whole day of conventional rehabilitation therapy.

## Discussion

In this study, we examined the safety, feasibility, and efficacy of wearable vibrotactile stimulation gloves and shirts for upper extremity stroke rehabilitation for patients during their stay in the rehabilitation unit. No participant experienced an adverse event related to the wearable devices, suggesting that the devices were safe. Treatment and control groups did not have significant differences in FMA-UE at admission (see Fig 5), which facilitated the comparison between groups at discharge. Both groups showed significant improvement in FMA-UE motor function from admission to discharge (see Fig 6A), but no significant difference was observed between groups (primary outcome). These within-group improvements did not extend to sensation, passive joint motion, or joint pain scores; although the treatment group exhibited a trend towards improvement (see Figs 6B and 6D). Prior work reported an improvement in sensation with vibrotactile stimulation (Schabrun and Hillier, 2009), which indicates that longer exposure times (both in duration or length) may have been needed to observe these improvements.

The only significant difference between groups was the FMA-UE joint pain change. This finding is clinically relevant because post-stroke upper-extremity pain is common after stroke, affecting approximately 22–47% of stroke survivors. The observed improvement in joint pain was associated with a large effect size (r = 0.53), supporting the potential clinical relevance of vibrotactile stimulation for reducing post-stroke upper-extremity pain. Beyond discomfort, joint pain can make it more difficult for stroke survivors to use their affected limb during therapy and everyday tasks, which may delay recovery and reduce quality of life (Lindgren et al., 2007;

Harrison and Field, 2015; Hettiarachchi et al., 2011; Ali et al., 2023). This is consistent with prior work in other disease states, including fibromyalgia, where vibrotactile stimulation has been shown to improve pain-related symptoms (Seim et al., 2023b; Staud et al., 2011; Pujol et al., 2019). These findings suggest that vibrotactile stimulation may help reduce post-stroke joint pain, which could facilitate greater participation in rehabilitation and use of the affected upper extremity. Because the assessments were conducted when patients were not wearing the devices, this indicates that vibrotactile stimulation has more than just an immediate effect reducing joint pain. For the secondary outcome, no significant difference in the total MAS score was observed between groups (see Fig 9). Although the unadjusted analysis suggested a significant between-group difference for the wrist MAS, this effect did not remain significant after correcting for multiple comparisons (see Fig 9C).

Responder analyses showed consistently higher responder rates in the treatment group than the control group for FMA-UE motor function, sensation, passive joint motion, and joint pain, with the latter outcome being statistically significant. For the MAS, responders were observed in the treatment group for both the fingers and wrist, whereas no responders were observed in the control group. While the differences did not reach significance, these findings suggest a possible effect of vibrotactile stimulation on upper-extremity spastic hypertonia; however, larger studies are needed to determine whether these preliminary trends represent true treatment effects.

No responders were observed for the elbow in both groups. One possible explanation is that the hand and wrist have dense sensory innervation and may therefore be more responsive targets to vibrotactile stimulation. Increased sensory input may help modulate spinal reflex pathways and reduce motor neuron excitability, contributing to reduced spasticity (Seim et al., 2023b). Future studies could investigate whether stimulating the impaired upper extremity (from the shoulder to the fingertips) yields additional benefits than stimulating the hand alone.

The duration and length of the vibrotactile stimulation treatment was likely too short to produce robust differences between groups. Prior work reporting significant improvements had chronic stroke patients wear vibrotactile stimulation gloves for three hours daily over eight weeks (Seim et al., 2021). Furthermore, our limited sample size (12 participants per group) may have reduced the ability to detect treatment effects. In future work, we plan to provide the devices for patients to take home, thereby enabling longer treatment durations and greater cumulative exposure to vibrotactile stimulation. Overall, our study suggests that some participants may benefit more than others from vibrotactile stimulation, which is particularly important in a small feasibility study where group-level effects may appear modest despite some meaningful individual responses.

Participants indicated that the wearable devices were well tolerated and expressed a desire to continue using them at home for their rehabilitation. However, sweating and mild itching during prolonged use were reported, suggesting limited breathability. Future designs should incorporate breathable and moisture-wicking materials, reduce material thickness, and optimize component placement to improve ventilation and user comfort. Eleven out of twelve participants completed the study in the treatment group.

We chose to apply the vibrotactile stimulation at a frequency range of 80-100 Hz, which corresponds to frequencies (and associated amplitudes) that lie on the lower bound of the range of values supported by our ERM motors. Prior work indicates that this frequency range, at low amplitudes, can activate muscle spindles that directly reaches primary somatosensory cortex and primary motor cortex (Heath et al., 1976; Huerta et al., 1990). This increased cortical excitability may engage plasticity mechanisms in spared sensory-motor circuits that improve motor recovery after stroke (Marconi et al., 2011). While the vibration motors on the shirts likely engaged muscle spindles, those on the fingertips likely engaged Pacinian corpuscles mechanoreceptors (Seim et al., 2021). Recent work from Seim and colleagues suggests that cutaneous stimulation at 250 Hz may be more effective at reducing spastic hypertonia than muscle spindle stimulation (Seim et al., 2023). Comparing to electrical stimulation, a common alternative non-invasive stimulation modality (Meyers et al., 2025), vibrotactile stimulation is a more natural form of afferent stimulation that may more effectively modulate excitability in motor cortical circuits (Rosenkranz et al., 2003). Determining the most effective stimulation modality (e.g., cutaneous, muscle spindle, electrical) and parameters (e.g., frequency, amplitude, location, and duration) to improve stroke rehabilitation outcomes is an open research problem.

Prior work with neck muscle vibration has shown beneficial effects for the treatment of hemineglect (Schindler et al., 2002; Stammler et al., 2026). In addition to neck muscle vibration, there is some preliminary evidence of beneficial effects of upper extremity muscle stimulation (Lucente et al., 2019). Here, we report some anecdotal evidence from one of our patients who had severe left neglect. The morning after the first session of vibrotactile stimulation, the therapist rolled the patient to her left side, and the patient said “my arm is vibrating. Do you feel that vibration? [no vibration was being applied]”. With her left neglect, she had made no comments about her left arm until that point. It would be valuable to investigate whether our stimulation shirts and gloves could constitute an effective treatment for hemineglect.

This study had several limitations. The FMA-UE and MAS scores depended on a rater’s judgement, which may have some variability despite attempts at standardization. There was no inclusion of a sham control in the study design, making it difficult to distinguish between the actual effects of vibrotactile stimulation compared to other non-specific effects like greater attention on the injured body part or psychological effects from the use of the device. This issue should be addressed in future research. An additional limitation of our study is the lack of follow-up assessments after patients are discharged from the rehabilitation unit. Prior work suggests that increases in sensorimotor cortex excitability from sensory stimulation in the subacute stroke period are correlated with motor recovery over the first year post-stroke (Schaechter et al., 2012). Marconi and colleagues showed that 3 consecutive days of 1-hour sessions of vibrotactile stimulation had long-term effects that lasted at least two weeks (Marconi et al., 2011). In follow-up studies, we plan to develop computer vision assessments of motor recovery that can be performed in the patient’s home to obtain longitudinal information after their discharge from the rehabilitation unit.

In conclusion, this clinical trial suggests that wearable vibrotactile stimulation devices are feasible, safe, and well-tolerated for post-stroke upper-extremity rehabilitation. The findings indicate that the most noticeable short-term benefits may be related to reducing joint pain, while potential effects on distal spasticity warrant further investigation. Effects on motor recovery and sensory perception may require longer intervention periods, larger sample sizes, or stimulation protocols combined with active rehabilitation. Future studies should focus on improving device comfort, extending treatment duration, and clarifying the mechanisms through which vibrotactile stimulation may influence pain, spasticity, and upper-limb recovery.

## Data Availability

All code and de-identified participant data used in this study, including the scripts for statistical analyses and figure generation can be found at the following address: https://github.com/DrWalaaAyyad/wearable-vibrotactile-stroke-rehabilitation. The full trial protocol, including the statistical analysis plan, is available in the Supporting Information (S3 Protocol).

## Acknowledgements

We would like to thank Al Borno Lab at the University of Colorado Anschutz Medical Campus for supplying our study with all necessary resources and materials. Special thanks to Jesse Gilmer, Gunnar Enserro, Madhuresh Gore, Ryan Priddy, Brody Skaufel, Kyle Rosen, Maab Taha, Sama Kasmani for investigating the system design. We also thank the OTs at UCHealth Rehabilitation Unit-University of Colorado Hospital and Broomfield Hospital for their cooperation and for conducting the FMA-UE and MAS for stroke patients included in this study. We are grateful to Dr. Joseph Rosenthal and Ashley Peter for facilitating the study at the rehabilitation unit.

## Authors’ contributions

Walaa Ayyad designed the study, developed the experimental protocol, coordinated participant recruitment, coordinated data collection, analyzed the data, and drafted the manuscript.

Yeongdae Kim contributed to the development of the wearable vibrotactile devices, assisted with the recruitment of initial study participants, and edited the manuscript.

Nathan Odom contributed to the study design, assisted with the recruitment of participants, and edited the manuscript.

Mazen Al Borno designed the study, contributed to the design and development of the wearable devices, supervised the research project, obtained Institutional Review Board (IRB) approval, provided guidance on data interpretation, and critically reviewed and revised the manuscript.

## Ethics approval and consent to participate

This study was approved by the Colorado Multiple Institutional Review Board (COMIRB Protocol #23-0783). Written informed consent was obtained from all participants.

## Supporting information

**S1 Fig.**
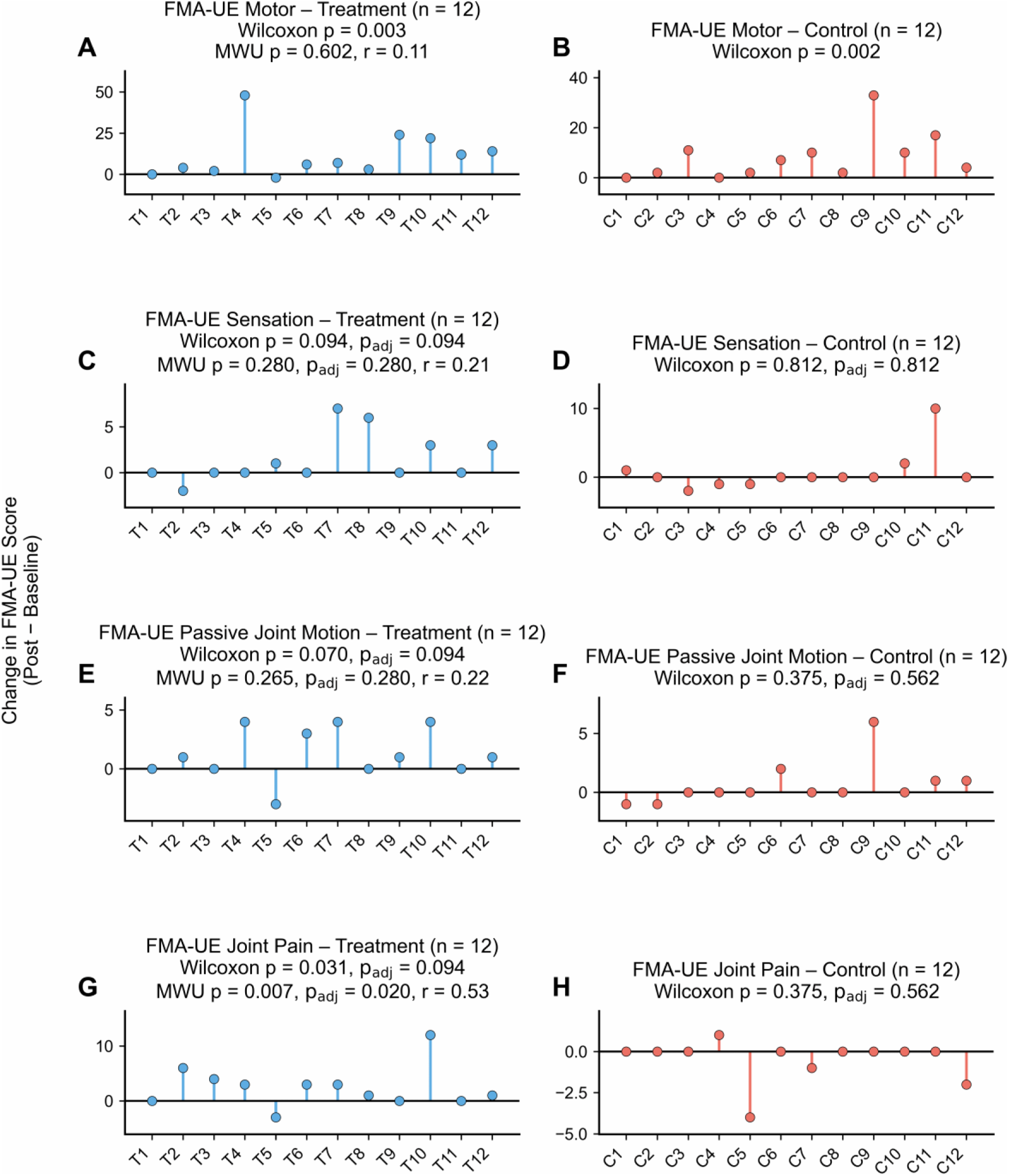
Individual FMA-UE change scores from admission to discharge. Each point represents an individual participant, and vertical lines connect the change score to zero. Panels A, C, E, and G show participants in the treatment group, while panels B, D, F, and H show participants in the control group. (A–B) Motor function, (C–D) sensation, (E–F) passive joint motion, and (G–H) joint pain. Significant within-group improvements were observed for motor function in both the treatment (p = 0.003) and control (p = 0.002) groups. Joint pain improved in the treatment group before, but not after, FDR correction (p = 0.031, p_adj_ = 0.094). Between-group differences were significant only for joint pain (p = 0.007, p_adj_ = 0.020, r = 0.53).

**S2 Fig.**
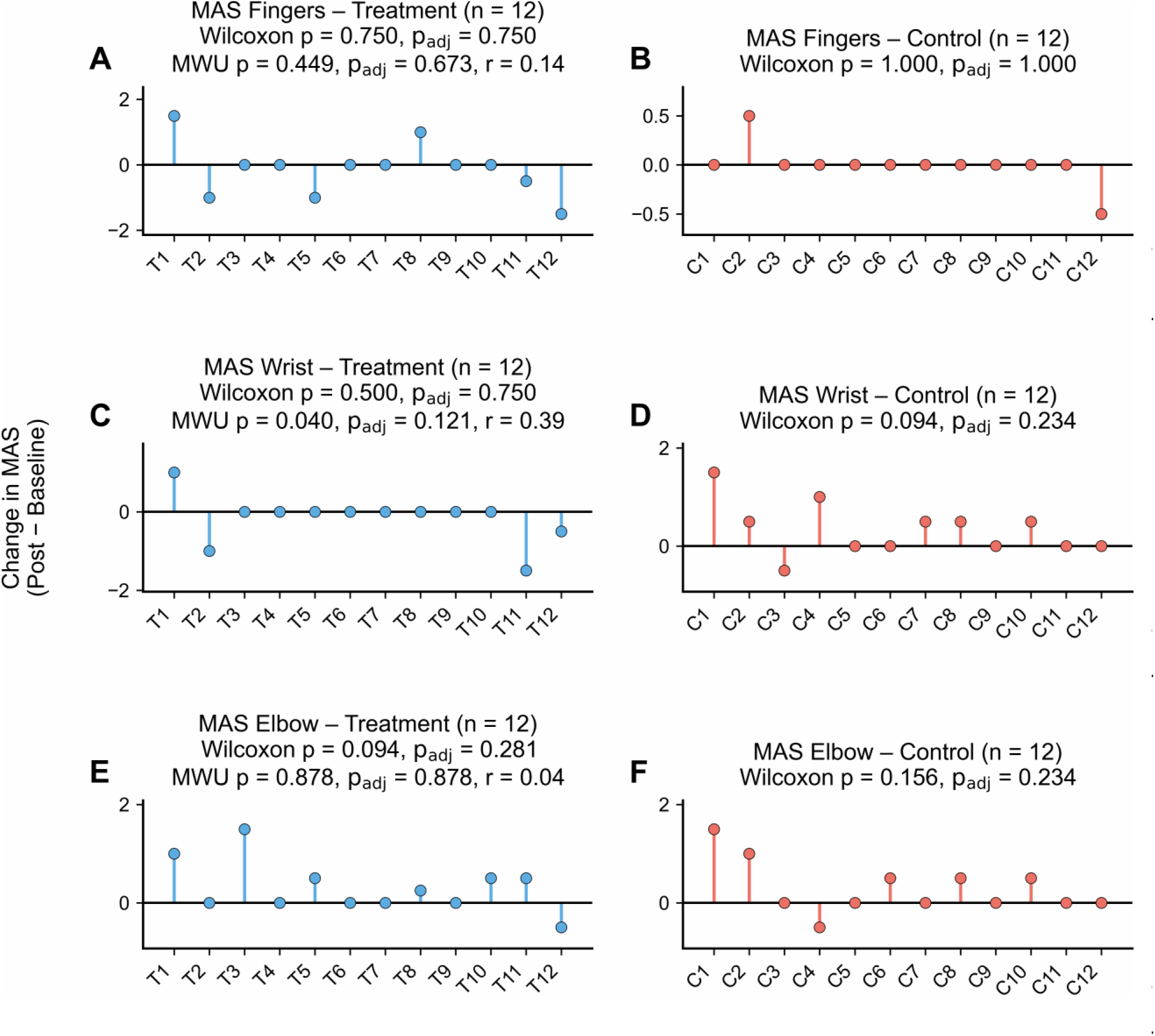
Individual MAS change scores from admission to discharge. Each point represents an individual participant, and vertical lines connect the change score to zero. Panels A, C, and E show participants in the treatment group, while panels B, D, and F show participants in the control group. (A–B) Fingers, (C–D) wrist, and (E–F) elbow. No significant within-group changes were observed for fingers, wrist, or elbow MAS scores in either group after FDR correction. Between-group differences were not significant for fingers (Mann–Whitney p = 0.449, p_adj_ = 0.673, r = 0.14), wrist (p = 0.040, p_adj_ = 0.121, r = 0.39), or elbow (p = 0.878, p_adj_ = 0.878, r = 0.04). Although the unadjusted between-group comparison for wrist MAS reached significance (p = 0.040), this effect did not remain significant after correction for multiple comparisons. MWU = Mann–Whitney U test.

